# TYK2 Inhibition with Deucravacitinib Improves Clinical Outcomes and Resolves Interferon-Driven Inflammation in Lichen Planopilaris

**DOI:** 10.64898/2026.03.13.26348272

**Authors:** Alyssa Stockard, Zachary Leibovit-Reiben, Bowen Hu, Rundong Jiang, Bryce T. Roberts, Scott Penner, Xing Li, Zeni Ramirez, Keegan Stewart, Jennifer Fox, Rachael Bogle, Nan Zhang, Samantha Shao, Henrique Borges Da Silva, David J. DiCaudo, Samantha Zunich, Thais P. Pincelli, Lam C. Tsoi, Mark R. Pittelkow, Jason Sluzevich, Johann E. Gudjonsson, Aaron R. Mangold

## Abstract

Lichen Planopilaris (LPP) is a lymphocyte-mediated scarring alopecia characterized by progressive follicular destruction and fibrosis. In this clinical trial, patients with biopsy proven LPP were treated with deucravacitinib (an oral inhibitor of tyrosine kinase 2 (TYK2)) 6 mg BID for 24 weeks (NCT-06091956). Bulk and single-cell RNA sequencing was performed on paired pre- and post-treatment scalp biopsies from baseline and week 4. Patients (N=10) demonstrated improvements in PGA (88.9%, p=0.008), LPPAI (−2.3 points, SD 1.1, p=0.002) and Skindex-16 (−21.0 points, SD 22.1, p=0.014) scores at week 24. Bulk transcriptomic analysis of untreated LPP revealed upregulation of type I Interferon (IFN)-stimulated genes and pathways related to inflammation, immune activation, keratinization, and extracellular matrix remodelling, with downregulation of immune and inflammatory pathways following treatment. Single-cell RNA-seq of LPP was characterized by enrichment of CD8+GZMK+ T cells which showed downregulation of T-cell receptor signaling as well as antiviral pathways with treatment. Basal keratinocytes exhibited reduced cytokine and interferon signaling and decreased communication with NK cells following treatment. CCL19+ fibroblasts were prominent in untreated disease was attenuated after treatment, with downregulation of type I IFN signaling. Selective TYK2 inhibition with deucravacitinib effectively suppresses these inflammatory circuits in LPP and represents a promising therapeutic strategy.

## Introduction

Lichen Planopilaris (LPP) is a form of lymphocyte-mediated scarring alopecia characterized by progressive follicular destruction and fibrosis. It presents discrete patches with characteristic perifollicular erythema and scale involving the scalp. LPP is histopathologically characterized by lichenoid inflammation at the infundibulum and isthmus of the hair follicle. Cell-mediated cytotoxicity is the main pathogenetic mechanism in lichenoid tissue reactions and the maintenance and progression of chronic cytotoxic inflammation requires Type I and II IFN signaling.^1–3^ Disease progression results in permanent alopecia and is associated with a negative impact on quality of life, including depression and anxiety.^4^ There are no approved therapies for LPP and conventional therapies display mixed responses.^5^

Cutaneous lichen planus (LP) is characterized by a Type I and Type II IFN driven cell mediated cytotoxic immune response that is highly responsive to JAK1, 2 inhibition.^1–3^ More moderate responses are seen with JAK1, 2 inhibition in LPP.^6^ Unlike LP, LPP is a chronic, scarring condition. Both the etiology and associated scarring of LPP are poorly understood, and there is potential contribution of Th17 cells.^6–8^ Taken together, we believe that TYK2 inhibition has the potential to treat the inflammation of LPP through Type I IFN blockade as well as the scarring generated by Th17 stimulated fibroblasts. Here, we report the results from a multi-center, exploratory, open-label, single-arm phase II clinical trial using a systemic TYK2 inhibitor for the treatment of LPP.

## Results

### Patients

11 participants were enrolled and 9 participants completed the study, with elective withdrawals prior to the week 2 and week 16 visits, respectively. 10 participants with follow-up data from multiple study visits were included in the analysis. The mean (SD) age at enrollment was 61.4 (11.7), with 70% female and 100% of patients identifying as White. The mean (SD) disease duration across all patients was 75.8 months (48.4). All patients had LPP refractory to prior therapy, with 100% failing topical steroids, 10% failing oral and intramuscular steroids, and 80% failing systemic treatments. The mean (SD) number of all treatments and systemic treatments per patient were 4.1 (1.8), and 1.7 (1.3), respectively. Demographics, treatment history, and outcomes for each patient are summarized in Table S1.

The mean (SD) disease activity by Lichen Planopilaris Activity Index (LPPAI) was 3.8 (1.2) at baseline. The overall mean (SD) baseline Dermatology Life Quality Index (DLQI) & Skindex-16 were 3.8 (2.0) and 36.3 (18.6), respectively. Baseline mean (SD) pruritus Numerical Rating Scale (NRS), average 24-hour pruritus Visual Analog Scale (VAS), and maximum 24-hour VAS scores were 4.2 (2.4), 3.4 (2.4), and 4.6 (3.1), respectively.

### Efficacy

At weeks 16 and 20, 9 patients (100%) demonstrated treatment response, with Physician Global Assessment (PGA) scores of 0 to 3, corresponding with ≥ 50% score reduction in disease activity (Figure 1A-B; Table S2). At week 24, 8 (88.9%) patients remained treatment responsive; 5 patients (55.6%) had a PGA of 2 (marked improvement) and 3 patients (33.3%) had a PGA of 3 (moderate improvement) (Figure 1C-D; Table S2). Improvement in PGA was observed on average at 7.2 weeks (SD 4.4). Treatment effects were sustained at week 28 (4 weeks off-therapy), with 7 (77.8%) patients demonstrating continued response by PGA score (Figure 1A-B; Table S2).

**Figure 1:**
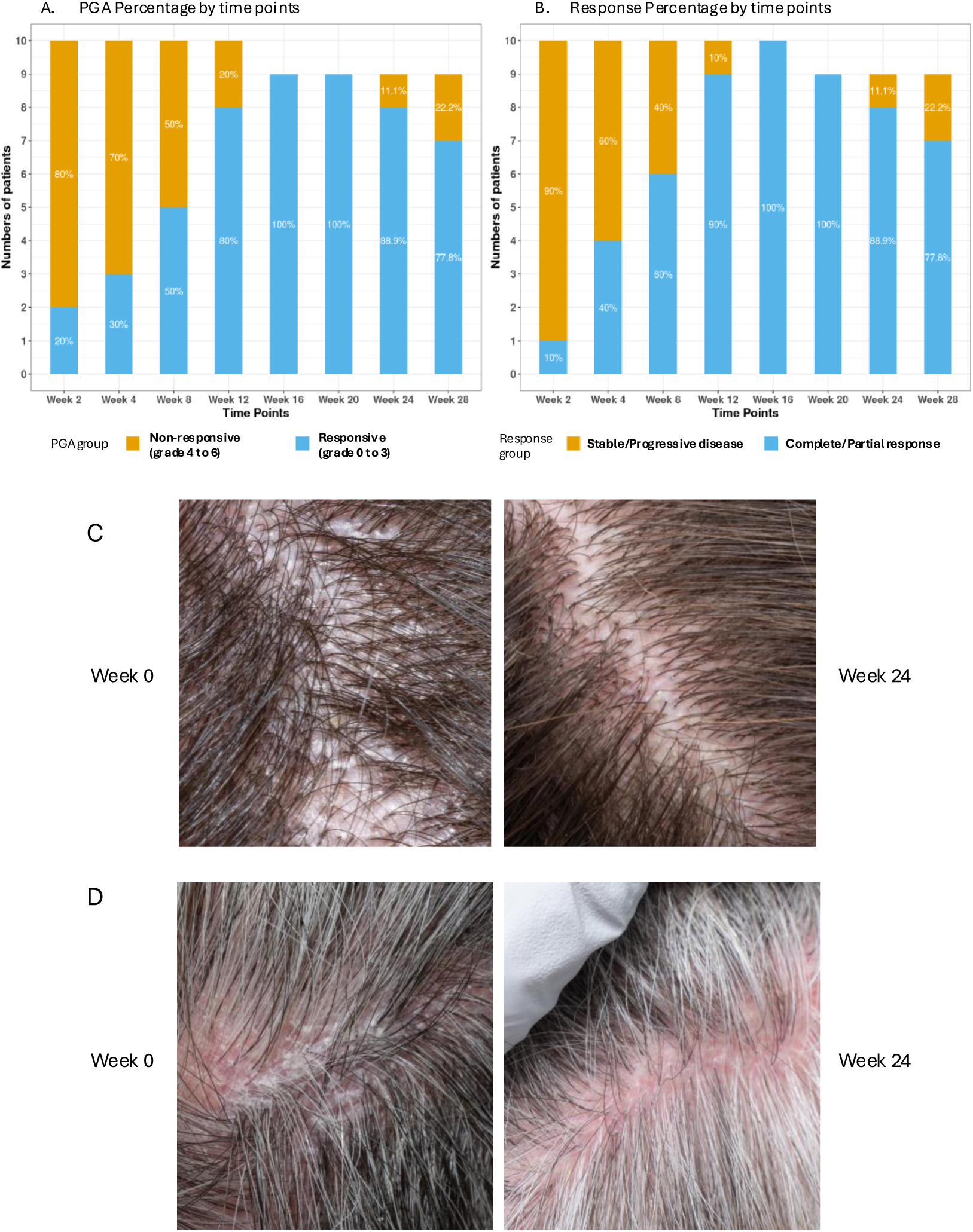
Primary Study Endpoint and Treatment Response. A. Physician Global Assessment (PGA) percentage by timepoints. B. Treatment response percentage by time points. Treatment response rates defined by PGA from baseline. C. Example image of PGA 2 (Marked Improvement). D. Example image of PGA 3 (Moderate Improvement).

Improvements were seen across multiple secondary measures at week 24, with statistically significant improvements in LPPAI and Skindex-16 scores (Table 1; Figure S1 A-D). The mean (SD) LPPAI decreased from a baseline of 3.8 (SD 1.2) to 1.5 (SD 1.1; p = 0.002). The decrease in itch and skin pain was non-significant (Table 1). Pruritus NRS improvement ≥4 from baseline (NRS4) was achieved in 33.3% of patients at week 24. The overall Skindex-16 score decreased from baseline to week 24 by a mean of –21.0 points (SD 22.1; p = 0.014) (Table 1), with decreases in each Skindex subscore: Symptom –3.6 (SD 7.2; p = 0.202), Emotional –14.3 (SD 13.1; p = 0.010), and Functional –3.1 (SD 5.7; p = 0.232). Results from the per-protocol analysis, with the population defined as patients who completed 24 weeks of deucravacitinib, were consistent with the results of the ITT analysis (Table S3).

**Table 1:** Primary and Secondary Endpoints at Baseline and Week 24 (ITT analysis)

|  | Baseline (n=10) | Week 24 (n=10) | Difference (n=10) | P value |
| --- | --- | --- | --- | --- |
| **PGA\*** |  |  |  | 0.034 |
| Responsive (n, %) | 2 (20%) | 8 (80%) |  |  |
| Non-responsive (n, %) | 8 (80%) | 2 (20%) |  |  |
| **LPPAI** |  |  |  | 0.002 |
| Mean (SD) | 3.8 (1.2) | 1.5 (1.3) | -2.3 (1.1) |  |
| Range | 1.7-5.5 | 0.3 - 4.1 | -4.2 - -1 |  |
| **NRS** |  |  |  | 0.175 |
| Mean (SD) | 4.2 (2.4) | 2.3 (1.9) | -1.9 (3.3) |  |
| Range | 1.0 - 7.0 | 1.0 - 7.0 | -6.0 - 5.0 |  |
| **Average VRS** |  |  |  | 0.018 |
| Mean (SD) | 1.5 (0.7) | 0.6 (0.5) | -0.9 (0.7) |  |
| Range | 1.0 - 3.0 | 0.0 - 1.0 | -2 - 0 |  |
| **Worst VRS** |  |  |  | 0.008 |
| Mean (SD) | 1.6 (0.7) | 0.7 (0.5) | -0.9 (0.6) |  |
| Range | 1.0 - 3.0 | 0.0 - 1.0 | -2 - 0 |  |
| **VAS - Avg 24 hr itch** |  |  |  | 0.131 |
| Mean (SD) | 3.4 (2.4) | 1.8 (1.8) | -1.6 (3.0) |  |
| Range | 0.0 - 6.6 | 0.2 - 6.1 | -5.2 - 4.7 |  |
| **VAS - Max 24 hr itch** |  |  |  | 0.064 |
| Mean (SD) | 4.6 (3.1) | 2.0 (1.9) | -2.7 (3.8) |  |
| Range | 1.0 - 10.0 | 0.1 - 6.3 | -8.9 - 5.1 |  |
| **Skindex-16** |  |  |  | 0.014 |
| Mean (SD) | 36.3 (18.6) | 15.3 (10.8) | -21.0 (22.1) |  |
| Range | 9.0 - 64.0 | 6.0 - 35.0 | -52.0 - 10.0 |  |
| **Skindex Symptom Subscore** |  |  |  | 0.202 |
| Mean (SD) | 7.7 (5.7) | 4.1 (2.4) | -3.6 (7.2) |  |
| Range | 1.0 - 16.0 | 1.0 - 9.0 | -14.0 - 8.0 |  |
| **Skindex Emotional Subscore** |  |  |  | 0.010 |
| Mean (SD) | 23.7 (11.7) | 9.4 (7.5) | -14.3 (13.1) |  |
| Range | 8.0 - 42.0 | 2.0 - 24.0 | -36.0 - 6.0 |  |
| **Skindex Functional Subscore** |  |  |  | 0.232 |
| Mean (SD) | 4.9 (5.7) | 1.8 (3.0) | -3.1 (5.7) |  |
| Range | 0.0 - 15.0 | 0.0 - 8.0 | -13.0 - 3.0 |  |
| **DLQI** |  |  |  | 0.629 |
| Mean (SD) | 3.8 (2.0) | 3.8 (4.0) | 0.0 (4.2) |  |
| Range | 2.0 - 8.0 | 1.0 - 13.0 | -7.0 - 10.0 |  |

### Safety

There were a total of 31 adverse events (AEs), with 13 deemed related to the study drug (Table S4, Table S5). These 13 AEs were mild, and acne was the most common AE (69.2%), affecting 9/10 patients (Table S5). Overall, deucravacitinib was well tolerated, with no drug related, serious treatment-emergent adverse events (TEAEs). Patient-level AEs are summarized in Table S5.

### Molecular Profiling of Lesional and Non-Lesional Tissue

Whole transcriptomic analysis using bulk RNA-seq was performed on lesional and healthy control scalp skin prior to therapy (n=7, 4, respectively). Differential expression analyses were conducted to identify differentially expressed genes (DEGs) (After False Discovery Rate (FDR) control, adjusted p value ≤ 0.05, and log_2_(Fold Change (FC)) ≥ 1). The differential analysis for lesional LPP skin vs. control samples at Day 0 revealed 1,782 DEGs, with 1,339 upregulated and 443 downregulated compared to unaffected control skin (Figure 2A). Among the most prominent upregulated DEGs in lesional LPP skin were IFN-stimulated genes MX1 (FC=1.32, FDR-adjusted p value = 0.0436), OAS2 (FC=1.91, p = 0.0001), IFI27, an IFN-inducible chemokine CXCL9 (FC=5.40, p = 0.009); chemokines involved in immune cell trafficking and lymphoid homing CXCL13 (FC=5.34, p = 0.0015), CCL19 (FC=2.09, p = 0.0047), CCR7 (FC = 3.10, p = 0.000003), and Type 1 inflammation/T-cell recruitment CCL5 (FC=3.78, p = 0.008); skin inflammatory cytokines IL36A (FC = 5.42, p = 0.047), IL36G (FC=2.34, p = 0.007), and fibrosis and extracellular matrix remodeling genes COL1A1 (FC=2.51, p = 0.001), COL1A2 (FC = 1.6, p = 0.004), COL3A1 (FC = 1.64, FDR = 0.029), COL5A1 (FC = 1.84, p = 0.003), MMP1 (FC = 5.18, p = 0.001), MMP3 (FC = 6.16, p = 0.012), MMP9 (FC = 2.89, p = 0.00002), MMP13 (FC = 9.08, p = 0.00003). Gene function/pathway enrichment analysis using the Gene Ontology database showed significant upregulation of pathways related to inflammation, immune activation, keratinization, and extracellular matrix remodeling (Figure 2B), and downregulation of cellular metabolic processes (Figure 2C). Taken together, baseline lesional LPP exhibited an interferon-enriched inflammatory signature with concurrent keratinization and extracellular matrix remodeling, indicating simultaneous immune activation and tissue remodeling with relative attenuation of metabolic processes.

**Figure 2:**
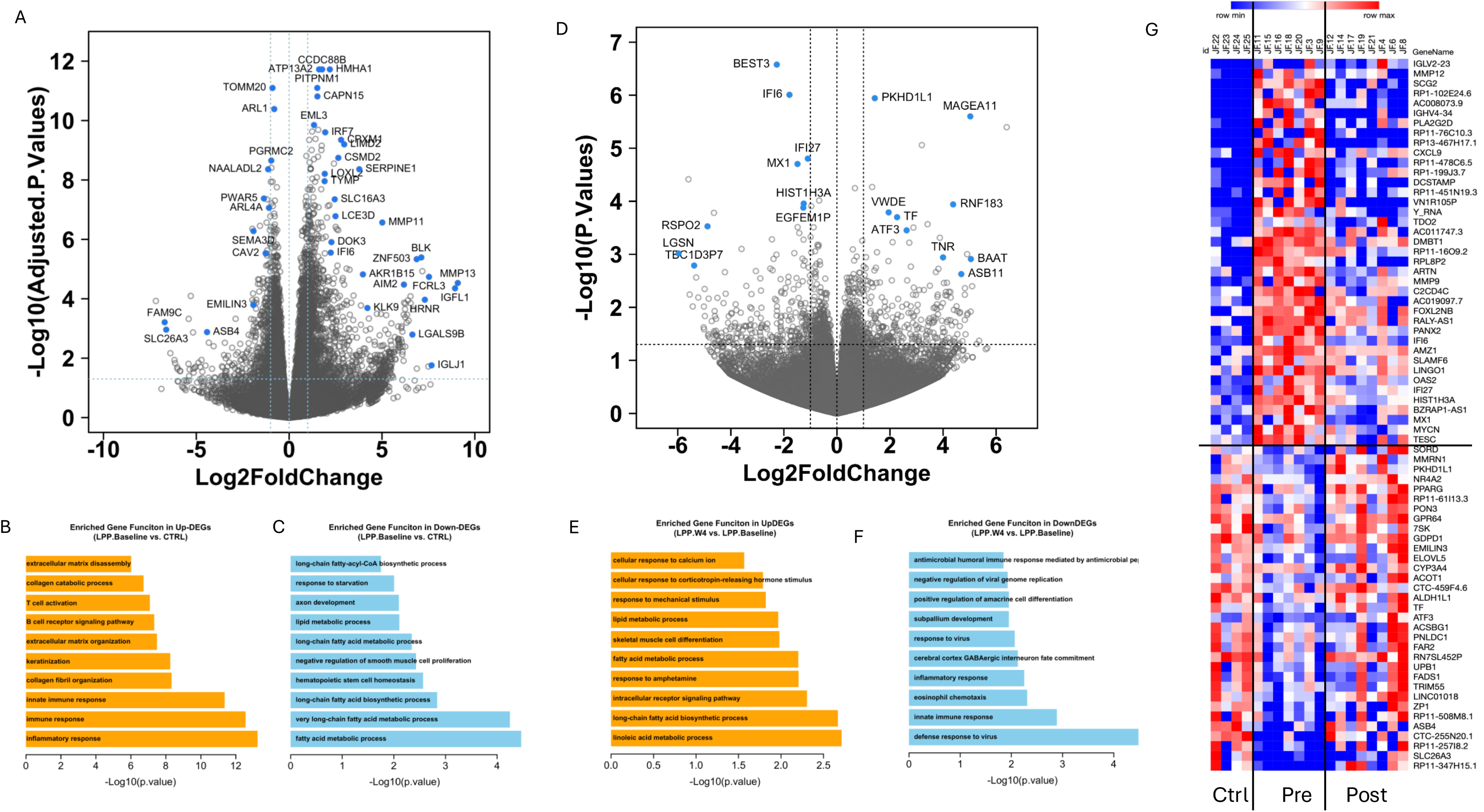
Transcriptomic Profiling and Treatment Effect of Deucravacitinib in Lichen Planopilaris. A. Volcano plot of bulk RNA-Seq data comparing lesional LPP versus control scalp at day 0 (n=7 and 4, respectively) with highly significant genes highlighted. B. Top 10 enriched pathways in the upregulated DEGs in untreated LPP. C. Top 10 enriched pathways in the downregulated DEGs in untreated LPP. D. Volcano plot of bulk RNA-Seq data comparing pre-versus post-treatment LPP at day 0 vs week 4 (n=7 and 8, respectively) with highly significant genes highlighted. E. Top 10 enriched pathways in the upregulated DEGs in post treatment LPP. F. Top 10 enriched pathways in the downregulated DEGs in post treatment LPP. G. Heatmap (Control n=4, Pre-treatment n=7, Post-treatment n=8) DEGs in disease baseline compared to control are reversed after treatment. Post-treatment gene expression profiles are more similar to control gene expression profiles.

### Molecular Profiling of Treatment Response

Whole transcriptomic analysis using bulk RNA-seq was performed on affected scalp tissues after 4 weeks of treatment with deucravacitinib and compared to pre-treatment lesional samples (n = 8, 7, respectively). Differential analysis using P value < 0.05 & |Log2FC| >1 for pre-versus post-treatment LPP skin revealed 531 DEGs, with 276 upregulated and 255 downregulated (Figure 2D). Gene set enrichment analysis showed downregulation of viral response, immune and inflammatory response pathways and upregulation of cellular metabolic pathways (Figure 2E-F). Downregulated fatty acid metabolism pathways and upregulated immune response pathways in pre-treatment LPP (Figure 2B-C) were reversed at 4-weeks post-treatment (Figure 2E-F).

### Disease gene expression profiles

By merging the differential expression gene lists between LPP disease baseline (pre-treatment) vs. control and post-treatment vs. pre-treatment, critical gene expression profiles downregulated or upregulated by treatment were identified (Figure S2A-B). Gene interaction network analysis was performed demonstrating an upregulated Interferon-rich cluster of hub genes at baseline, including *OAS2*, *IFI6*, *IFI27, CXCL9* and *MX1,* and tissue remodeling genes including *MMP9* and *MMP12* (Figure S2A). The downregulated hub-genes in disease baseline included *PPARG*, *ALDH1L1*, *ACSBG1*, *ELOVL5*, *CYP3A4*, and *SORD* (Figure S2B). Treatment normalized gene expression differences in the disease tissue.

### Single Cell Transcriptional Profile of LPP-associated cells

To better assess the cellular mechanisms involved in LPP, we performed scRNA-seq from baseline and week 4 lesional LPP biopsies from 10 patients. After quality control (see Methods), we identified 69,250 cells with 230–2,269 genes (median 661) and 220–3,320 transcripts (median 739) detected per cell (10th–90th percentiles), from 8 affected donors with PGA scores of 2, 3, and 5 at week 0, paired post-treatment lesions at week 4, and 5 unaffected donors. Using the unbiased clustering method from Seurat R package (v5.0.1), we identified 28 cell clusters and overlapped these with known canonical cell type markers to annotate 15 major cell types (Figure 3A, C). All 15 major cell types were identified in all three groups of patient responders, with treatment-associated shifts in multiple cell populations with the largest transcriptional changes observed in keratinocytes, L endothelial cells, and lymphocytes (Figure 3B, D, E). To understand how keratinocytes contribute to the pathogenesis of LPP and changes in their function during treatment, we subclustered keratinocytes into basal, cycling, and differentiated, and keratinized keratinocytes (Figure 3A). Basal and differentiated keratinocytes were the predominant keratinocyte subtypes in control samples, while cycling keratinocytes were most abundant in pre-treatment LPP samples (Figure 3C-D). Data after 4 weeks of treatment showed a reduced proportion of cycling keratinocytes and >150 DEGs in basal keratinocytes between pre- and post-treatment samples (Figure 3D-E). Basal keratinocytes exhibited reduced cytokine and interferon signaling post-treatment (Figure 4A).

**Figure 3:**
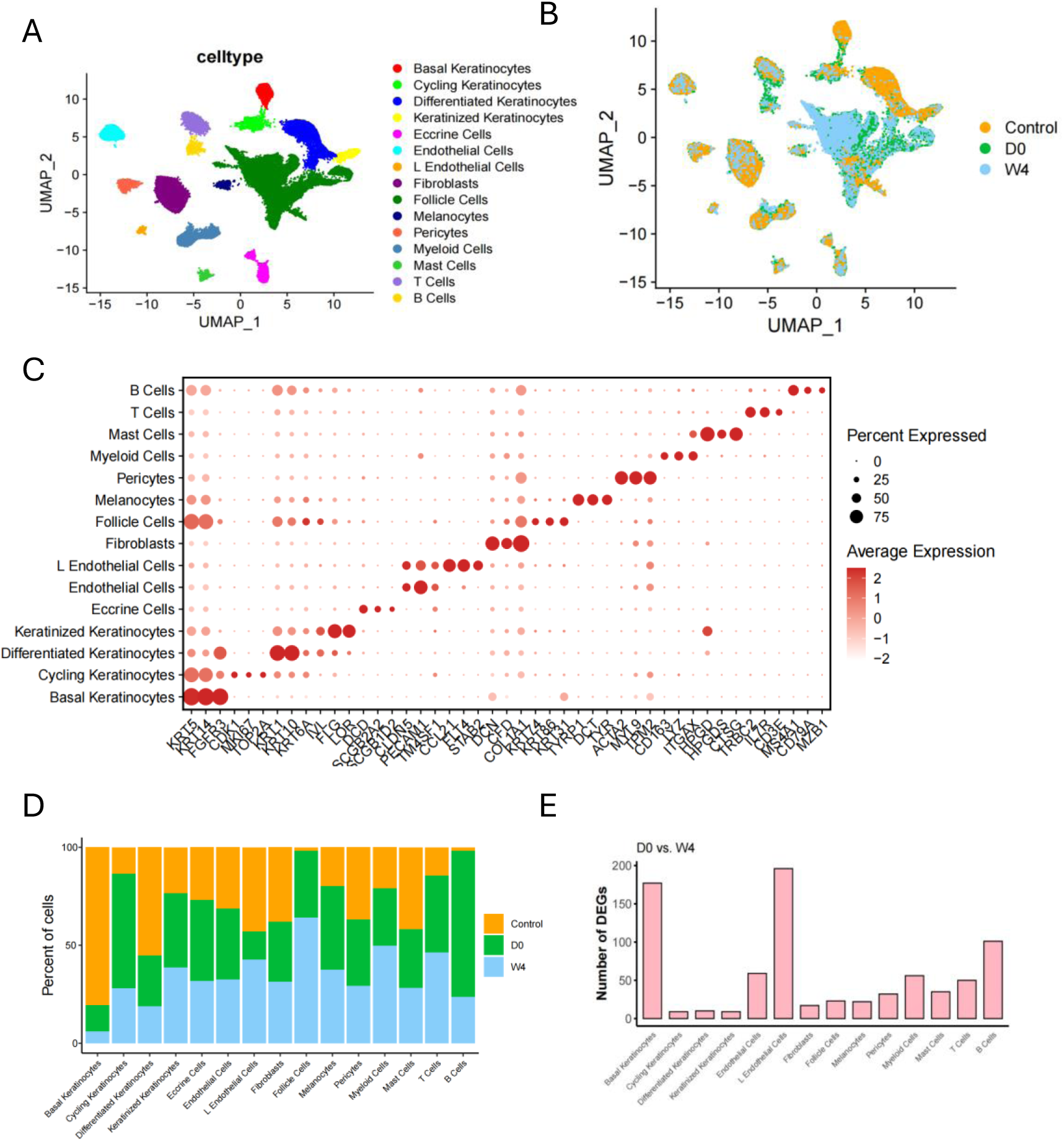
Single Cell Composition of Lichen Planopilaris. A. Integrated UMAP defines 15 cell clusters colored by major cell class. B. Integrated UMAP colored by control, baseline (day 0) and week 4 (n = 5, 8, 8, respectively). C. Dot plot showing the scaled expression of marker genes in major cell types identified in lesional LPP skin. D. Bar graph showing proportion of control, day 0, and week 4 across the major cell types. E. Bar graph showing the number of DEGs day 0 vs week 4 across the major cell types.

**Figure 4:**
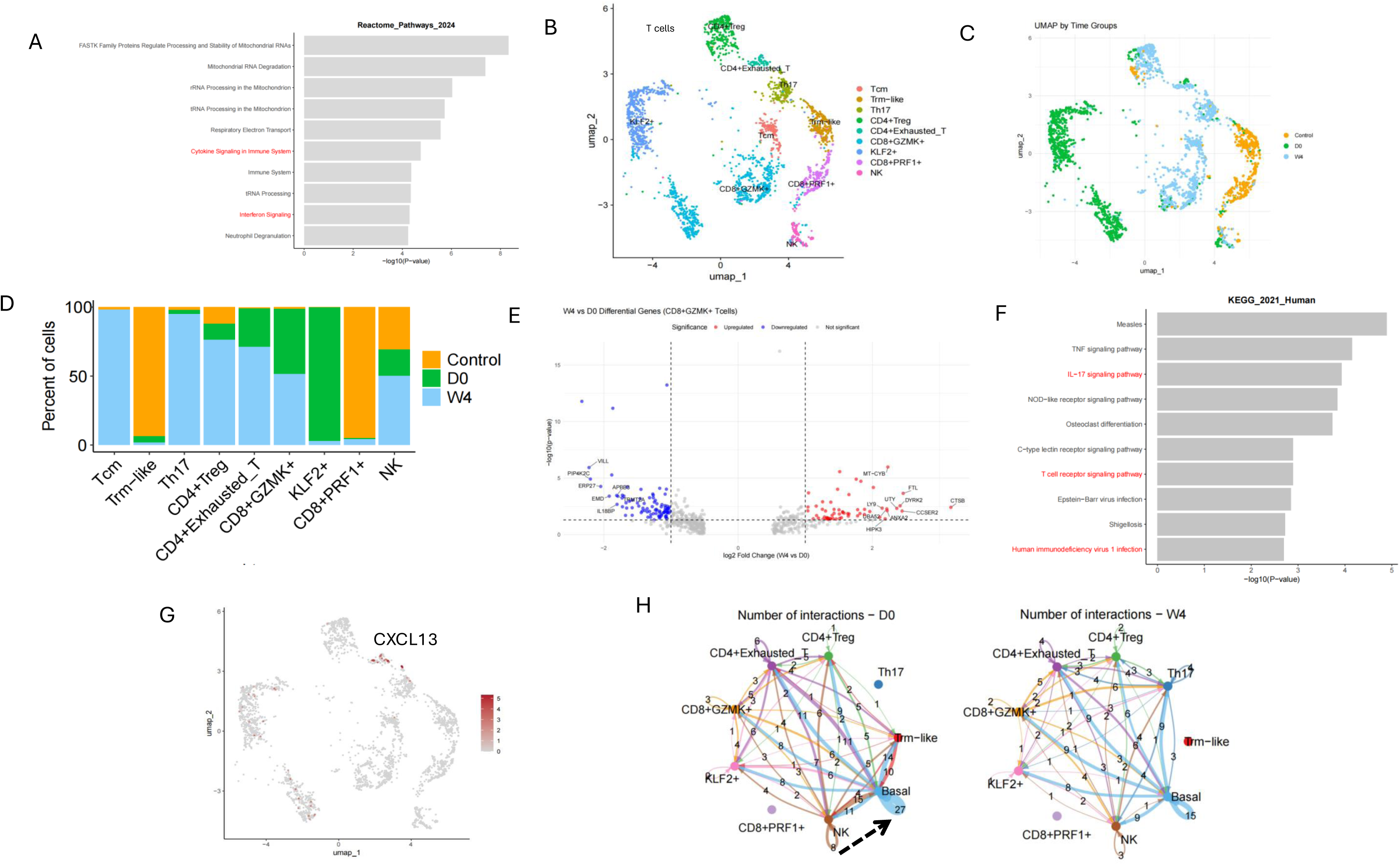
Lymphocyte Profile and Keratinocyte Interactions. A. Reactome Top 10 enriched pathways of downregulated differentially expressed genes in basilar keratinocytes. B. Nine lymphocyte subsets are found in LPP skin. C. Lymphocyte subsets by control, day 0, and week 4 (n = 5, 8, 8, respectively). D. Lymphocyte proportions for control, day 0, and week 4. E. Volcano plot of differentially expressed genes of CD8+GZMK+ T-cells. F. KEG top 10 enriched pathways of downregulated differentially expressed genes in CD8+GZMK+ T-cells with treatment. G. CXCL13 expression. H. CellChat analysis demonstrating decreased communication between NK cells and basilar keratinocytes.

LPP has prominent perifollicular T-cell infiltration, but the nature of the T-cell involvement has not previously been addressed. We identified 9 subclusters of T-cells in LPP, including Tcm, Trm-like, Th17, CD4+ Treg, CD4+ exhausted, *CD8*+*GZMK*+, *KLF2*+, *CD8*+*PRF1*+, and NK T cells (Figure 4B). Untreated LPP was characterized by enrichment of KLF2+ and CD8+GZMK+ T cells (Figure 4C-D). With deucravacitinib treatment, CD8+GZMK+ T-cells showed downregulation of IL-17 signaling, T-cell receptor signaling, and antiviral pathways (Figure 4E-F). CD8+GZMK+ T cells were the major source of IFNG in pre-treatment samples, while CD8+CXCL13+ T-cells were absent in both pre- and post-treatment samples (Figure S3, Figure 4G). Pathway analysis of DEGs identified in NK cells demonstrated downregulation of IL-17 signaling and antiviral pathways, while basal keratinocytes exhibited reduced cytokine and interferon signaling post-treatment (Figure S4 A-B, Figure 4A). Notably, cell-cell communication analysis revealed decreased signaling between NK cells and basal keratinocytes following treatment (Figure 4H).

Fibroblasts play a key role in scar formation, although the nature of fibroblasts in the scarring of LPP is poorly understood. We identified 8 major fibroblast subsets (*CCL19+*, *COL11A1+*, *COL23A1+*, *CYP4B1+*, *LSP1+*, *SFRP2+*, *SFRP4+*, and *TNC*+) with predominant expression of *CCL19+* fibroblasts in D0 samples (Figure 5A-C). At week 4 there was a reduction of *CCL19+* fibroblasts compared to pre-treatment (Figure 5C). These *CCL19+* fibroblasts displayed a hallmark inflammatory response which was diminished after treatment (Figure 5D-E). Pathway enrichment analysis of DEGs in CCL19+ fibroblasts demonstrated downregulation of immune pathways and type I IFN signaling (Figure 5F-G).

**Figure 5:**
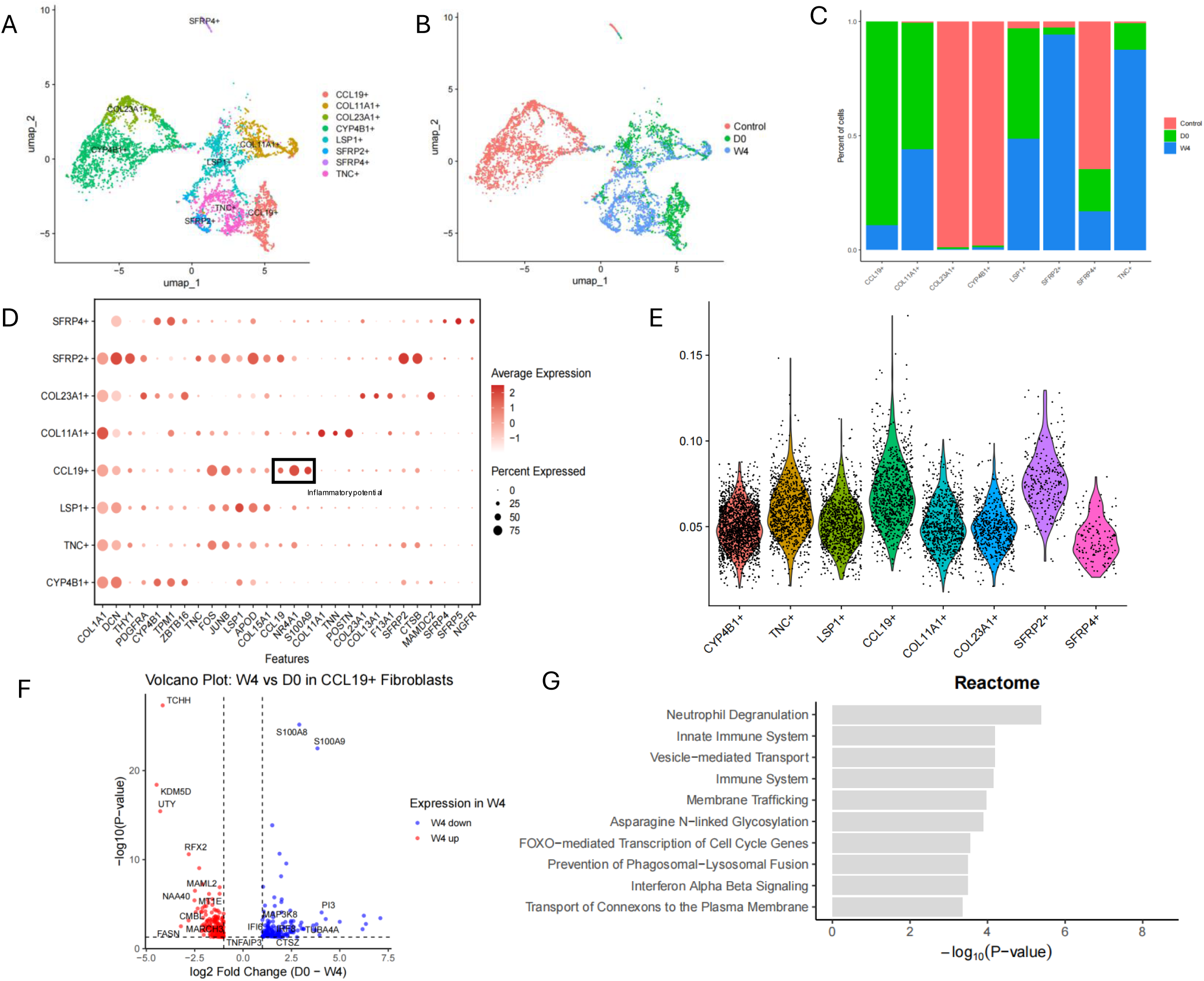
Single Cell Profile of Fibroblasts in Lichen Planopilaris. A. Eight fibroblast subsets are found in LPP skin. B. Fibroblast subsets by control, day 0, and week 4 (n = 5, 8, 8, respectively). C. Fibroblast proportions for control, day 0, and week 4. D. Marker genes in fibroblast cell types in LPP skin. E. Violin plot of Hallmark inflammation response (AUC score). F. Volcano plot of differentially expressed genes of CCL19+ Fibroblasts week 4 versus day 0. G. Reactome Top 10 enriched pathways of downregulated DEG in CCL19+ fibroblasts with treatment.

## Discussion

This open-label, single-arm study suggests that selective TYK2 inhibition with deucravacitinib may provide meaningful and sustained clinical benefit in patients with LPP, including those with chronic treatment-refractory disease. The high proportion of patients achieving clinically significant improvement across multiple outcome measures supports a potentially robust therapeutic effect. Notably, clinical responses were maintained in a substantial subset of patients following treatment discontinuation, indicating the deucravacitinib may induce durable modulation of disease activity rather than transient suppression alone. These findings are particularly relevant given the limited treatment options for LPP and the often-refractory nature of the disease. The observed response rates are comparable to those reported with brepocitinib, a dual JAK1/TYK2 inhibitor, suggesting that selective TYK2 inhibition may be sufficient to achieve clinically meaningful improvement while potentially offering a more targeted mechanisms of action.^9^

The pathogenesis of LPP is poorly understood. Prior research in LP and LPP has highlighted the central role of TH1 response, Type II IFN, and fibrosis, however, this data was largely focused on frontal fibrosing alopecia (FFA) and generalized scarring alopecia (FFA, LPP, and central centrifugal cicatricial alopecia).^9–11^ In classic LP, both Type I and II IFN play a central role in the disease trigger and propagation.^12,13^ In LPP, however, our transcriptomics analysis highlights a predominantly type I IFN milieu in untreated disease along with immune activation and tissue remodeling. Treatment response was associated with broad downregulation of immune and inflammatory response pathways following treatment. PPARG was a downregulated hub gene in untreated LPP and demonstrated increased expression with treatment. In vivo, deletion of PPARG has been shown to produce an LPP like phenotype in follicular stem cells in mice.^14^ Restoration of PPARG, which regulated cell sheath function, may aid in reducing fibrosis and scaring.^15^ Matrix metalloproteinases, which are involved with tissue remodeling and fibrosis, were downregulated with treatment highlighting the potential to decrease in inflammation and scarring in LPP.

We were also able to characterize the single cell profile of LPP in the context of selective TYK2 inhibition. Our single-cell RNA-seq identified treatment-associated shifts in multiple cell populations with the largest transcriptional changes observed in keratinocytes, L endothelial cells, and lymphocytes. Untreated LPP was characterized by enrichment of KLF2+ and CD8+GZMK+ T cells. KLF2+ T cell enrichment and increased CCR7 expression suggests a recirculating lymphoid program rather than tissue residency.^16^ This is supported by single cell comparisons of LP, LPP, and mucosal LP suggesting limited involvement of tissue-resident markers in LPP (Jiang R, manuscript accepted, pending publication Nat Comm). With deucravacitinib treatment, CD8+GZMK+ T-cells showed downregulation of T-cell receptor signaling as well as antiviral pathways. CD8+GZMK+ T cells have been associated with chronic, proinflammatory activity autoimmune diseases Sjogren’s syndrome, rheumatoid arthritis, SLE, and systemic sclerosis.^17,18^ Treatment with deucravacitinib downregulated IL-17 signaling-associated genes in CD8+GZMK+ T-cells. Perifollicular fibrotic LPP tissues has previously been shown to express IL-17 suggesting a potential role of CD8+GZMK+ T-cells in driving both inflammation and scarring.^7^ Notably, we did not observe CD8+CXCL 13+ T cells in LPP, which have been previously shown as the major source of IFNG expression in lesional LP skin, suggesting there are distinct processes active in LPP which differentiate it from LP.^13^

Basal keratinocytes exhibited reduced cytokine and interferon signaling on pathway analysis, as well as decreased communication between NK cells and basal keratinocytes following treatment. This data is consistent with other studies in lichenoid skin conditions which suggest interferon-mediated signaling drives communication between immune cells and keratinocytes in affected skin.^12,13^

Fibroblasts are a critical cell type in scarring and fibrosis, and our transcriptomic data revealed several fibroblast subtypes in LPP. In particular, inflammatory CCL19+ fibroblasts were prominent in untreated disease and displayed a hallmark inflammatory response, which was improved after treatment, with downregulation of type I IFN signaling. CCL19+ fibroblasts have been recently shown to drive fibrosis and dermal immune niches in systemic scleroderma.^19^ Our data suggests CCL19+ fibroblasts may play a role in the fibrosis and scarring of LPP.

This is the first study examining deucravacitinib in LPP, and the results demonstrate improvements in both disease activity and the effect of deucravacitinib on patient quality of life (QoL). Together, these findings indicate that LPP is driven by a type I IFN-dominant inflammatory program involving CD8+GZMK+ T cells, inflammatory CCL19+ fibroblasts and pathogenic immune-epithelial crosstalk. Selective TYK2 inhibition with deucravacitinib effectively suppresses these inflammatory circuits, providing mechanistic insights into therapeutic response and supporting TYK2 targeting as a promising therapeutic strategy for LPP.

## Methods

### Trial Design

This single-arm, open-label, phase 2, first-in-human trial was conducted at Mayo Clinic, Arizona and Florida (ClinicalTrials.gov identifier NCT-06091956). Ten patients with biopsy-proven cutaneous LP were administered oral deucravacitinib 6 mg BID for 24-weeks. The primary endpoint was an overall response by PGA of skin at week 24, with treatment response defined as PGA 0 to 3 (with ≥ 50% score reduction, Supplemental Table S1). Secondary outcomes were changes in LPPAI, DLQI, average 24-hour pruritus VAS, maximum 24-hour pruritus VAS, pruritus NRS, and Skindex-16.^20–23^ Patients were evaluated at baseline (week 0) and weeks 2, 4, 8, 12, 16, 20, 24, and 28.

Treatment efficacy, AEs, and QoL were assessed at each study visit. Patient scalps were evaluated, photographed, and scored using PGA and LPPAI scores. All clinical assessments were performed by the principal investigator and co-investigators (ARM, JS, SZ) with questionable assessments scored by 2 investigators. The revised National Cancer Institute Common Terminology Criteria for Adverse Events version 5.0 was used for AE reporting. We used intention-to-treat (ITT) analysis for early discontinuation with the patient’s last observation to impute week 24 data.

Tissue samples via 4 mm punch biopsies were collected at baseline and week 4 of LPP lesional skin and normal appearing skin for bulk RNA-sequencing. Week 4 samples were designated as responsive (defined as a lesion with ≥ 50% response by PGA grade 0-3) or nonresponsive (defined as a lesion with < 50% response by PGA grade 4,5). Standard photos were used at Week 0 and Week 4 for lesion identification; biopsies were taken at least 1 cm apart, and all biopsies were taken from the same lesion if possible, or from the same body region. Tissue samples were formalin-fixed and paraffin embedded (FFPE).

### Eligibility Criteria

Patients aged ≥ 18 years with biopsy-proven LPP were eligible for the trial. Both treatment-naïve and treatment-refractory disease were included. Key exclusion criteria included predominantly non-classical LPP (e.g. FFA), active infections, and other active inflammatory cutaneous conditions. See supplemental for additional eligibility criteria (Supplemental Note).

### Statistical Analysis

Patient demographics, clinical characteristics, and outcomes were summarized as mean, standard deviation, median, interquartile range for continuous variables, and frequency and percentages for categorical variables. Primary and secondary outcome differences between baseline and week 24 were compared using Wilcoxon signed rank test for continuous variables and McNemar’s test for binary variables. Exact binomial method was used to calculate the treatment response rate at week 24 and its corresponding 95% confidence interval. All analyses were conducted with R version 4.1.2 (R Foundation for Statistical Computing, Vienna, Austria). The cut-off for statistical significance was set as 0.05.

### Tissue processing, transcriptomic processing, quality control, alignment, and analyses

#### Bulk RNA-seq analysis

For bulk RNA-seq, tissue was processed, RNA was isolated as previously described by our group, and 150bp paired-end reads were generated.^24^ The reads were adapter trimmed and aligned to the human genome hg38, with only the uniquely mapped reads used for expression level quantification. DESeq2 was used to perform read normalization and differential expression analyses.

### Generation of single-cell suspensions for scRNA-seq

#### scRNA-seq data analysis

Data processing, including quality control, read alignment (hg38), and gene quantification, was conducted using the 10X Cell Ranger. The samples were then merged into a single expression matrix using the cellranger aggr pipeline. The R package Seurat (v3.1.2) was used to cluster the cells in the merged matrix.^25^ Cells with less than 500 transcripts or 100 genes, or more than 10% of mitochondrial expression, were first filtered out as low-quality cells. SoupX was utilized to remove ambient RNA reads. Doublets were detected and removed using scDblfinder.^26,27^ The NormalizeData function was used to normalize the expression level for each cell with default parameters. The FindVariableFeatures function was used to select variable genes with default parameters. The FindIntegrationAnchors and IntegrateData functions were used to integrate the samples prepared using different 10X Chromium chemistries. Samples were batch-corrected using Harmony, utilizing the donor as a batch. Sub-clustering was performed on the abundant T-cell types. The FindClusters function in the Seurat R package was used to obtain the sub-clusters. Sub-clusters defined exclusively by mitochondrial gene expression, indicating low quality, were removed from further analysis. Subtypes were annotated by overlapping sub-cluster marker genes with canonical subtype signature genes. Using CellChat (v2.1.2), we used the default computeCommunProb ‘trimean’ method, which approximates 25% truncated mean to calculate the average gene expression per cell group.^28^

## Study approval

The Mayo Clinic Institutional Review Board approved the study (IRB 23-001929), and all patients provided written informed consent prior to participation. All patients provided written informed consent for photographs, and the record of informed consent has been retained.

## Data availability

The RNA-Seq data discussed in this publication will be available in the NCBI’s Gene Expression Omnibus database under accession number *** at the time of publication.

## Acknowledgments

Funding for this study was provided by Bristol Myers Squibb. JEG and LCT are supported by NIH P30 AR075043.

## Author Contributions

Conceptualization: ARM, JEG

Protocol Writing: ARM

Data Collection: KS, SZ, TPP, JS, ARM

Statistical Analysis: NZ, XL

Tissue processing and Transcriptomic Analysis: BH, RJ, AH, XL, ZR,

Supervision: DJD, TPP, MRP, JS, JEG, HBdS, ARM

Visualization: BH, RJ, XL, NZ, SS

Writing - Original Draft Preparation: ALS, ZLR, XL, ARM, JEG

Writing – Review and Edit: ALS, ZLR, BH, RJ, BTR, SP, XL, ZR, KS, AH, NZ, DJD, SZ, TPP, MRP, JS, JEG, SS, HBdS, ARM

---

Supplementary Figure S1: Secondary Measures

A. LPPAI score by study time points.
B. NRS score by study time points.
C. Skindex-16 score by study time points.
D. DLQI score by study time points.

Supplementary Figure S2: Disease Gene Expression Profiles are Reversed After Treatment

A. Gene interaction network upregulated in LPP disease.
B. Gene interaction network downregulated in LPP.

Supplementary Figure S3: T-cell Expression of Key Cytotoxic Mediators

A. IFNG expression by control, day 0, and week 4.
B. PRF1 expression by control, day 0, and week 4.
C. GZMB expression by control, day 0, and week 4.

Supplementary Figure S4: Natural Killer Cells

A. Volcano plot of differentially expressed genes of NK T-cells.
B. KEG top 10 enriched pathways of downregulated differentially expressed genes in NK T-cells with treatment.

Supplementary Table S1: Patient Demographics, Baseline Disease Characteristics, and Outcomes

tCCs: topical corticosteroids, tAF: topical antifungals, tRUX: topical ruxolitinib, ilCCs: intralesional corticosteroids, oCCs: oral corticosteroids, oABX: oral antibiotics, MTX: methotrexate, HCQ: hydroxychloroquine, MPA: mycophenolate, 5-ARI: 5-alpha-reductase inhibitors (finasteride, deutasteride), BARI: baricitinib, oCNI: oral calcineurin inhibitors, ISO: isotretinoin, PRP: platelet-rich plasma, WD: Withdrew from trial.

Supplementary Table S2: Physician Global Assessment (PGA) Scores by Week

Supplementary Table S3 - Primary and Secondary Endpoints at Baseline and Week 24 (Per-Protocol Analysis)

Supplementary Table S4: Adverse Events

Table S5: Patient Level Adverse Events

