## Supplementary Figures for "TYK2 Inhibition with Deucravacitinib Improves Clinical Outcomes and Resolves Interferon-Driven Inflammation in Lichen Planopilaris"

### Slide 1
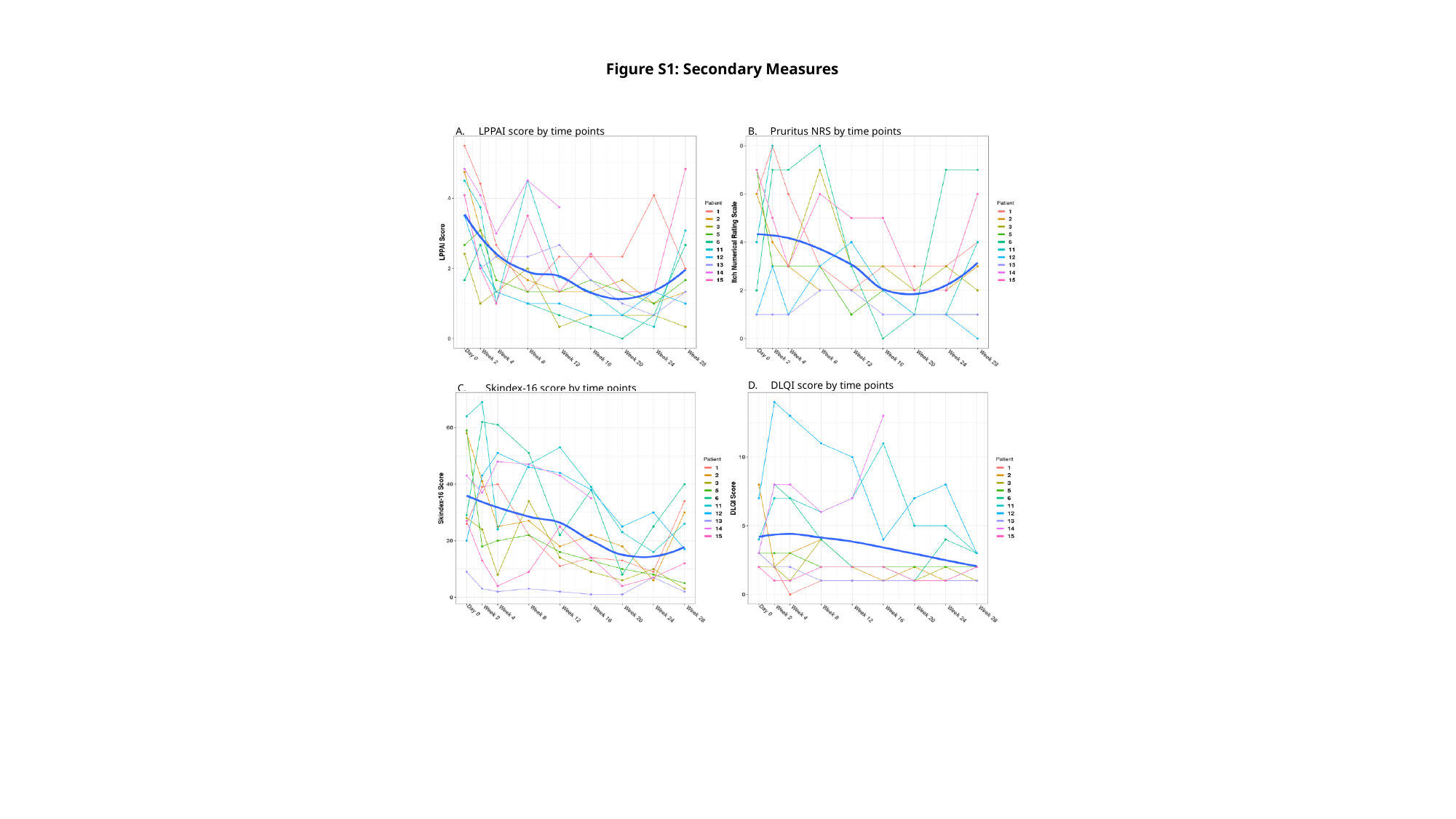

Figure S1: Secondary Measures
B.  Pruritus NRS by time points
LPPAI score by time points
D.  DLQI score by time points
C. Skindex-16 score by time points

### Slide 2
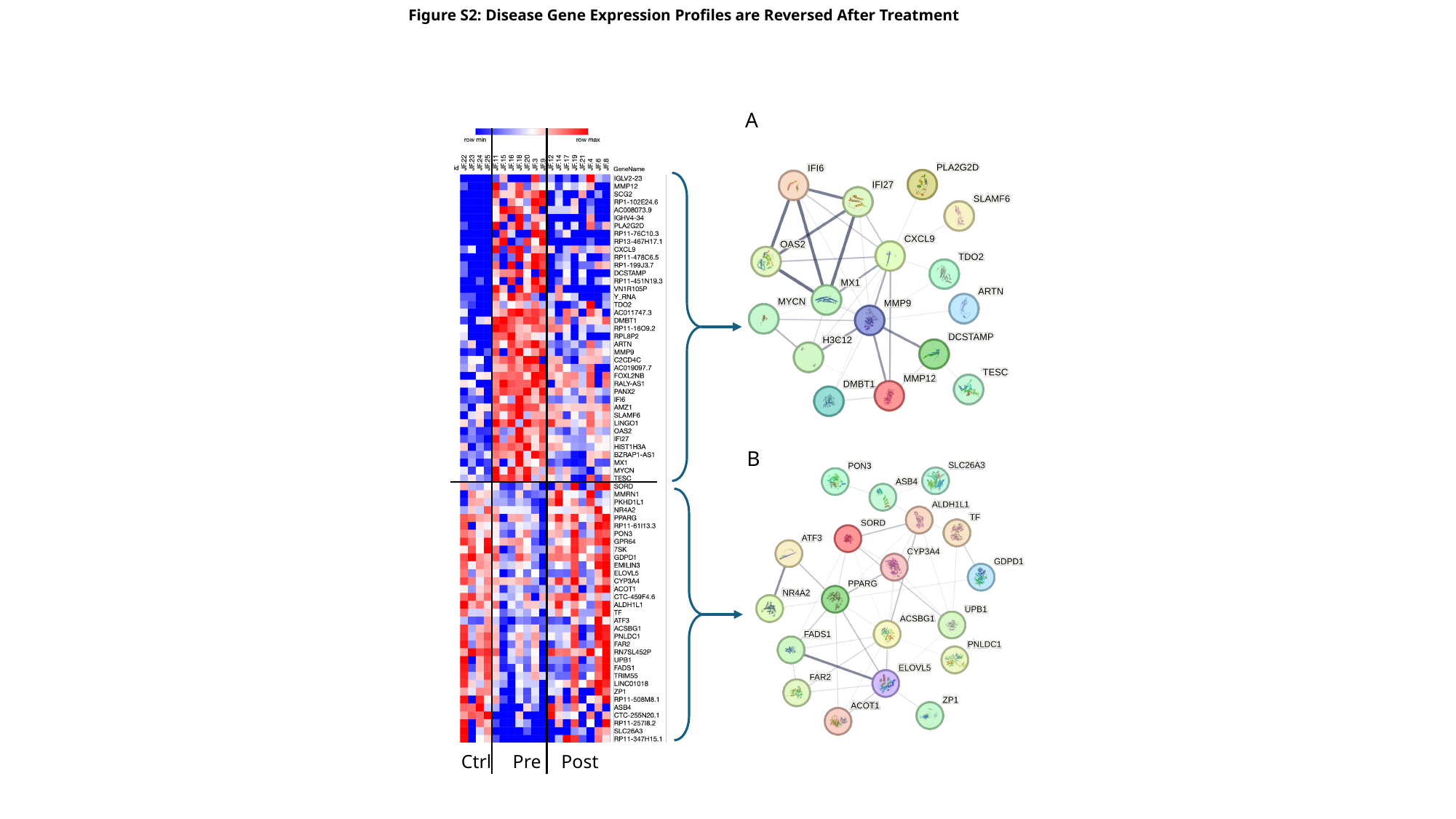

Figure S2: Disease Gene Expression Profiles are Reversed After Treatment
A
Ctrl
Pre
Post
B

### Slide 3
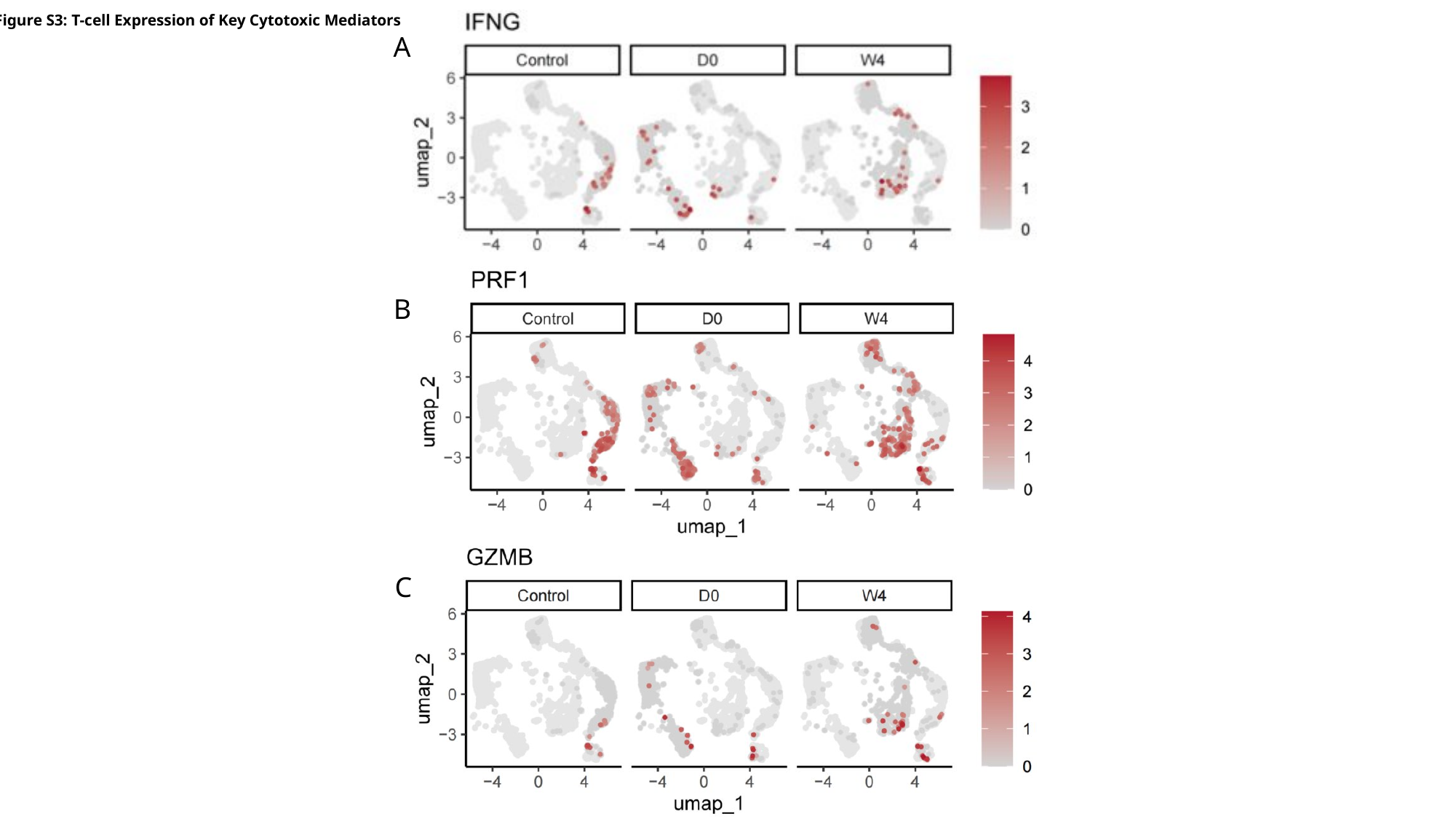

Figure S3: T-cell Expression of Key Cytotoxic Mediators
A
B
C

### Slide 4
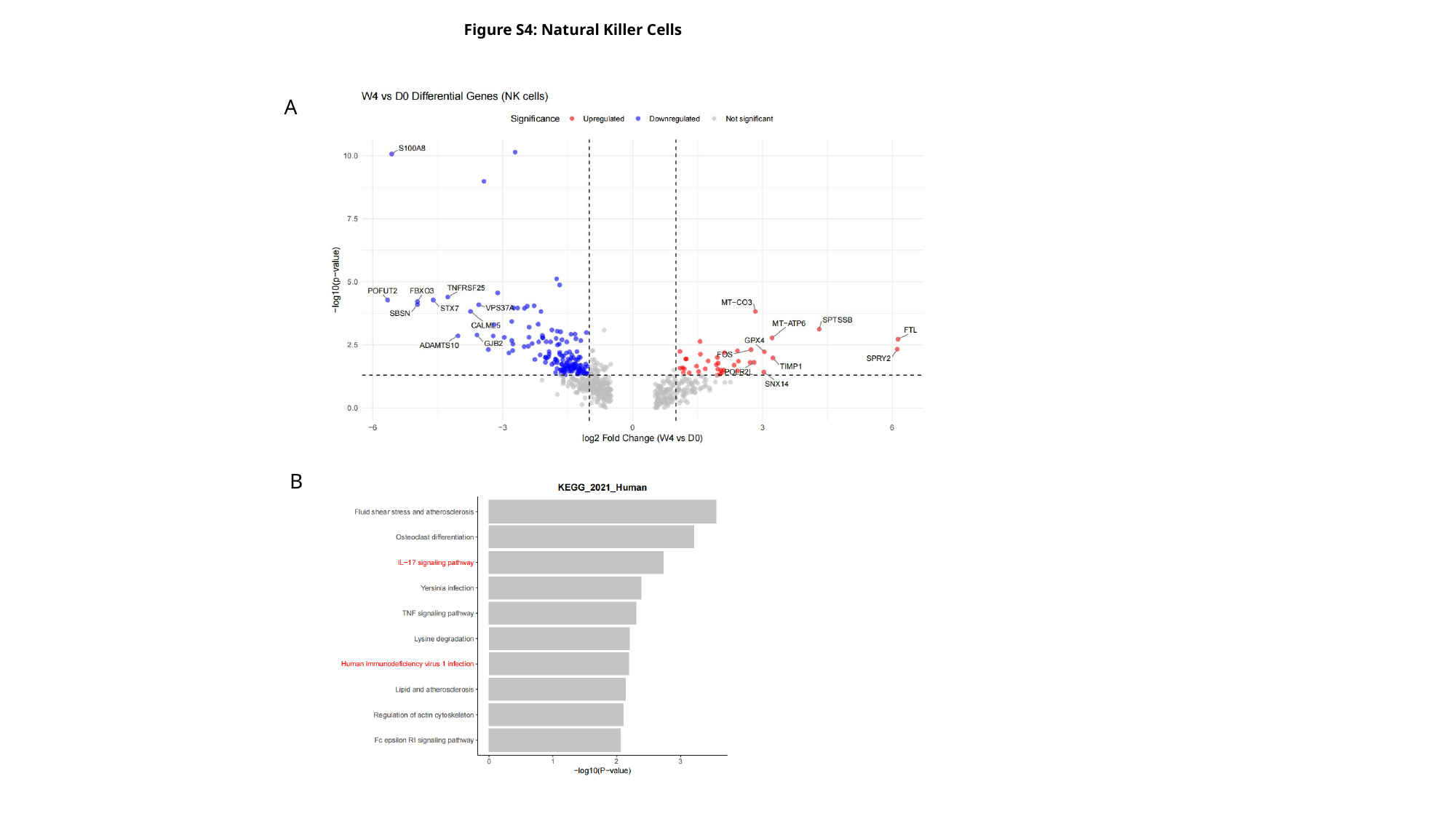

Figure S4: Natural Killer Cells
A
B

### Slide 5
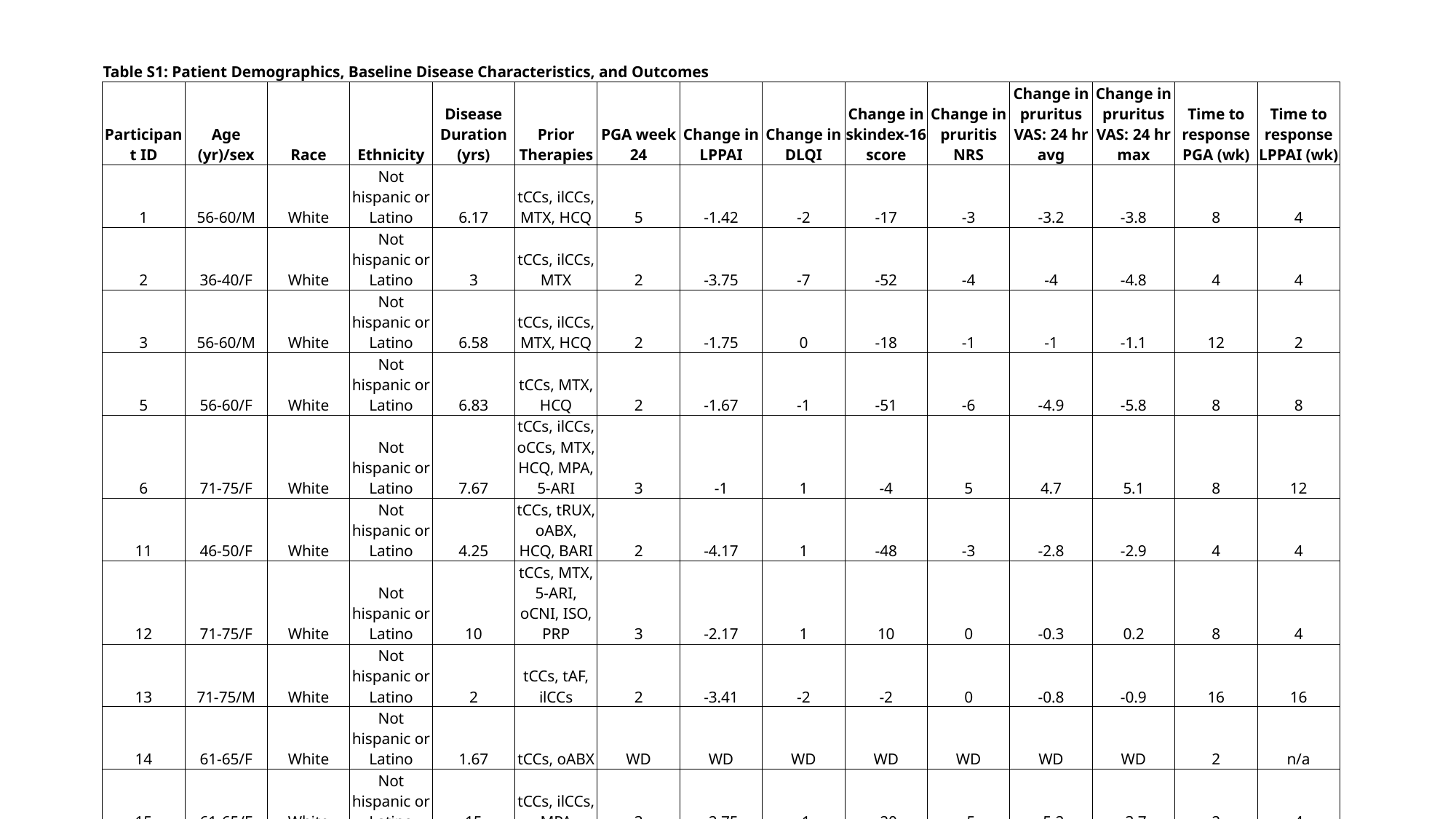

| Table S1: Patient Demographics, Baseline Disease Characteristics, and Outcomes | | | | | | | | | | | | | | |
| --- | --- | --- | --- | --- | --- | --- | --- | --- | --- | --- | --- | --- | --- | --- |
| Participant ID | Age (yr)/sex | Race | Ethnicity | Disease Duration (yrs) | Prior Therapies | PGA week 24 | Change in LPPAI | Change in DLQI | Change in skindex-16 score | Change in pruritis NRS | Change in pruritus VAS: 24 hr avg | Change in pruritus VAS: 24 hr max | Time to response PGA (wk) | Time to response LPPAI (wk) |
| 1 | 56-60/M | White | Not hispanic or Latino | 6.17 | tCCs, ilCCs, MTX, HCQ | 5 | -1.42 | -2 | -17 | -3 | -3.2 | -3.8 | 8 | 4 |
| 2 | 36-40/F | White | Not hispanic or Latino | 3 | tCCs, ilCCs, MTX | 2 | -3.75 | -7 | -52 | -4 | -4 | -4.8 | 4 | 4 |
| 3 | 56-60/M | White | Not hispanic or Latino | 6.58 | tCCs, ilCCs, MTX, HCQ | 2 | -1.75 | 0 | -18 | -1 | -1 | -1.1 | 12 | 2 |
| 5 | 56-60/F | White | Not hispanic or Latino | 6.83 | tCCs, MTX, HCQ | 2 | -1.67 | -1 | -51 | -6 | -4.9 | -5.8 | 8 | 8 |
| 6 | 71-75/F | White | Not hispanic or Latino | 7.67 | tCCs, ilCCs, oCCs, MTX, HCQ, MPA, 5-ARI | 3 | -1 | 1 | -4 | 5 | 4.7 | 5.1 | 8 | 12 |
| 11 | 46-50/F | White | Not hispanic or Latino | 4.25 | tCCs, tRUX, oABX, HCQ, BARI | 2 | -4.17 | 1 | -48 | -3 | -2.8 | -2.9 | 4 | 4 |
| 12 | 71-75/F | White | Not hispanic or Latino | 10 | tCCs, MTX, 5-ARI, oCNI, ISO, PRP | 3 | -2.17 | 1 | 10 | 0 | -0.3 | 0.2 | 8 | 4 |
| 13 | 71-75/M | White | Not hispanic or Latino | 2 | tCCs, tAF, ilCCs | 2 | -3.41 | -2 | -2 | 0 | -0.8 | -0.9 | 16 | 16 |
| 14 | 61-65/F | White | Not hispanic or Latino | 1.67 | tCCs, oABX | WD | WD | WD | WD | WD | WD | WD | 2 | n/a |
| 15 | 61-65/F | White | Not hispanic or Latino | 15 | tCCs, ilCCs, MPA | 3 | -2.75 | -1 | -20 | -5 | -5.2 | -3.7 | 2 | 4 |
| \*Age Range Reported to Limit Identifying Information | | | | | | | | | | | | | | |

### Slide 6
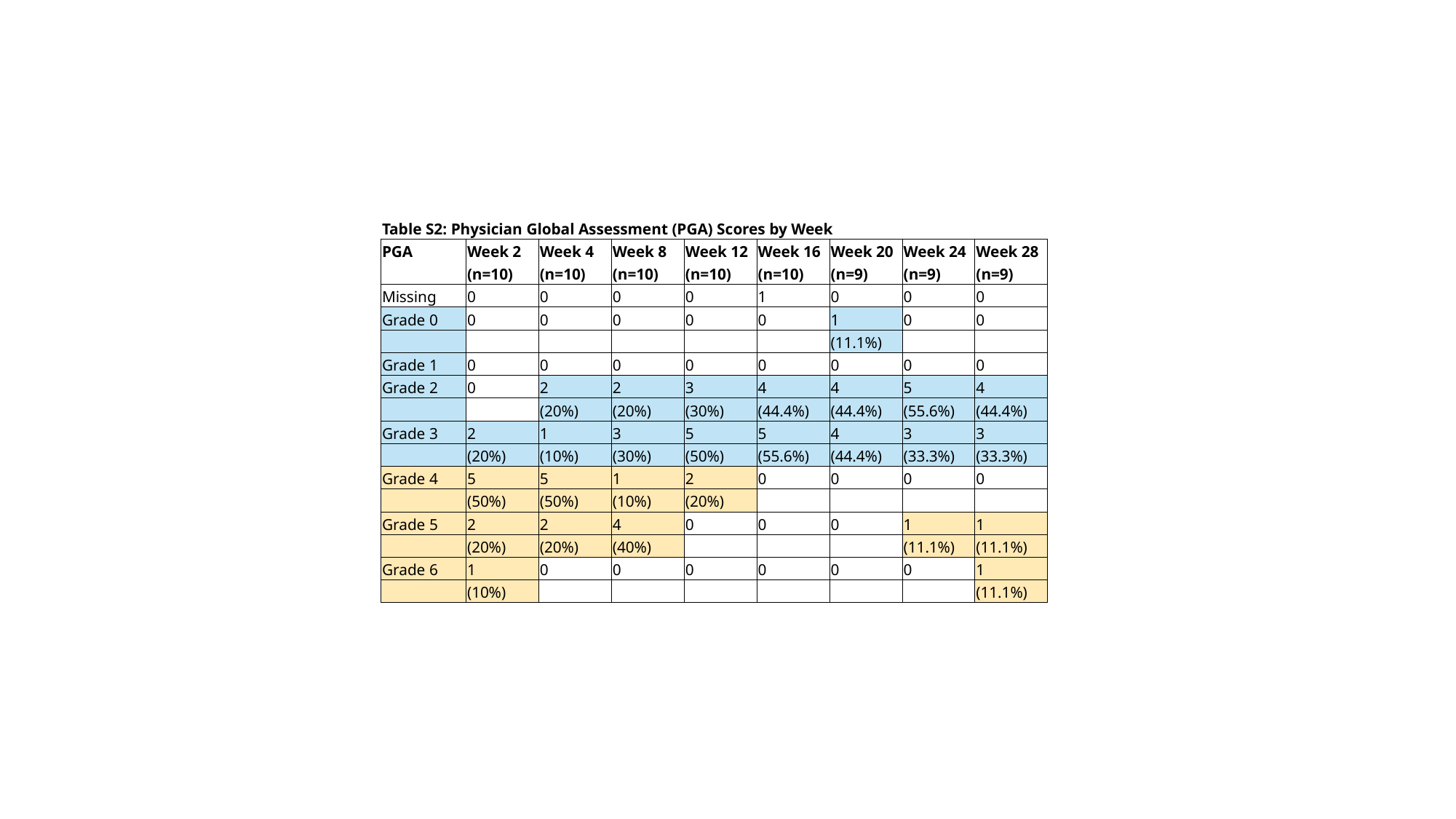

| Table S2: Physician Global Assessment (PGA) Scores by Week | | | | | | | | |
| --- | --- | --- | --- | --- | --- | --- | --- | --- |
| PGA​ | Week 2​ | Week 4​ | Week 8​ | Week 12​ | Week 16​ | Week 20​ | Week 24​ | Week 28​ |
| | (n=10)​ | (n=10)​ | (n=10)​ | (n=10)​ | (n=10)​ | (n=9)​ | (n=9)​ | (n=9)​ |
| Missing​ | 0​ | 0​ | 0​ | 0​ | 1​ | 0​ | 0​ | 0​ |
| Grade 0​ | 0​ | 0​ | 0​ | 0​ | 0​ | 1 ​ | 0​ | 0​ |
| | | | | | | (11.1%)​ | | |
| Grade 1​ | 0​ | 0​ | 0​ | 0​ | 0​ | 0​ | 0​ | 0​ |
| Grade 2​ | 0​ | 2 ​ | 2 ​ | 3 ​ | 4 ​ | 4 ​ | 5 ​ | 4 ​ |
| | | (20%)​ | (20%)​ | (30%)​ | (44.4%)​ | (44.4%)​ | (55.6%)​ | (44.4%)​ |
| Grade 3​ | 2 ​ | 1 ​ | 3 ​ | 5 ​ | 5 ​ | 4 ​ | 3 ​ | 3 ​ |
| | (20%)​ | (10%)​ | (30%)​ | (50%)​ | (55.6%)​ | (44.4%)​ | (33.3%)​ | (33.3%)​ |
| Grade 4​ | 5 ​ | 5 ​ | 1 ​ | 2 ​ | 0​ | 0​ | 0​ | 0​ |
| | (50%)​ | (50%)​ | (10%)​ | (20%)​ | | | | |
| Grade 5​ | 2 ​ | 2 ​ | 4 ​ | 0​ | 0​ | 0​ | 1 ​ | 1 ​ |
| | (20%)​ | (20%)​ | (40%)​ | | | | (11.1%)​ | (11.1%)​ |
| Grade 6​ | 1 ​ | 0​ | 0​ | 0​ | 0​ | 0​ | 0​ | 1 ​ |
| | (10%)​ | | | | | | | (11.1%)​ |

### Slide 7
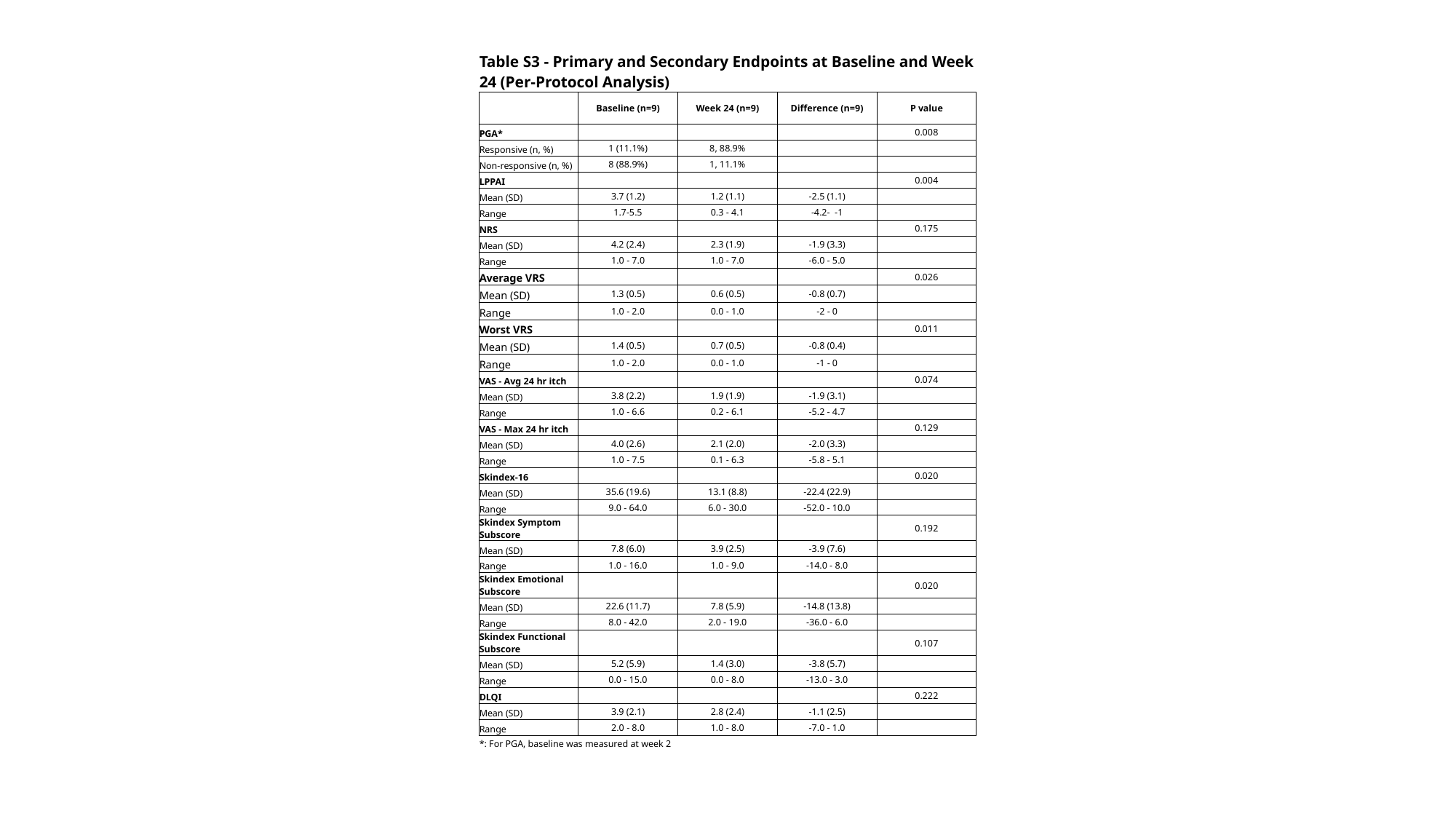

| Table S3 - Primary and Secondary Endpoints at Baseline and Week 24 (Per-Protocol Analysis) | | | | |
| --- | --- | --- | --- | --- |
| | Baseline (n=9) | Week 24 (n=9) | Difference (n=9) | P value |
| PGA\* | | | | 0.008 |
| Responsive (n, %) | 1 (11.1%) | 8, 88.9% | | |
| Non-responsive (n, %) | 8 (88.9%) | 1, 11.1% | | |
| LPPAI | | | | 0.004 |
| Mean (SD) | 3.7 (1.2) | 1.2 (1.1) | -2.5 (1.1) | |
| Range | 1.7-5.5 | 0.3 - 4.1 | -4.2- -1 | |
| NRS | | | | 0.175 |
| Mean (SD) | 4.2 (2.4) | 2.3 (1.9) | -1.9 (3.3) | |
| Range | 1.0 - 7.0 | 1.0 - 7.0 | -6.0 - 5.0 | |
| Average VRS | | | | 0.026 |
| Mean (SD) | 1.3 (0.5) | 0.6 (0.5) | -0.8 (0.7) | |
| Range | 1.0 - 2.0 | 0.0 - 1.0 | -2 - 0 | |
| Worst VRS | | | | 0.011 |
| Mean (SD) | 1.4 (0.5) | 0.7 (0.5) | -0.8 (0.4) | |
| Range | 1.0 - 2.0 | 0.0 - 1.0 | -1 - 0 | |
| VAS - Avg 24 hr itch | | | | 0.074 |
| Mean (SD) | 3.8 (2.2) | 1.9 (1.9) | -1.9 (3.1) | |
| Range | 1.0 - 6.6 | 0.2 - 6.1 | -5.2 - 4.7 | |
| VAS - Max 24 hr itch | | | | 0.129 |
| Mean (SD) | 4.0 (2.6) | 2.1 (2.0) | -2.0 (3.3) | |
| Range | 1.0 - 7.5 | 0.1 - 6.3 | -5.8 - 5.1 | |
| Skindex-16 | | | | 0.020 |
| Mean (SD) | 35.6 (19.6) | 13.1 (8.8) | -22.4 (22.9) | |
| Range | 9.0 - 64.0 | 6.0 - 30.0 | -52.0 - 10.0 | |
| Skindex Symptom Subscore | | | | 0.192 |
| Mean (SD) | 7.8 (6.0) | 3.9 (2.5) | -3.9 (7.6) | |
| Range | 1.0 - 16.0 | 1.0 - 9.0 | -14.0 - 8.0 | |
| Skindex Emotional Subscore | | | | 0.020 |
| Mean (SD) | 22.6 (11.7) | 7.8 (5.9) | -14.8 (13.8) | |
| Range | 8.0 - 42.0 | 2.0 - 19.0 | -36.0 - 6.0 | |
| Skindex Functional Subscore | | | | 0.107 |
| Mean (SD) | 5.2 (5.9) | 1.4 (3.0) | -3.8 (5.7) | |
| Range | 0.0 - 15.0 | 0.0 - 8.0 | -13.0 - 3.0 | |
| DLQI | | | | 0.222 |
| Mean (SD) | 3.9 (2.1) | 2.8 (2.4) | -1.1 (2.5) | |
| Range | 2.0 - 8.0 | 1.0 - 8.0 | -7.0 - 1.0 | |
| \*: For PGA, baseline was measured at week 2 | | | | |

### Slide 8
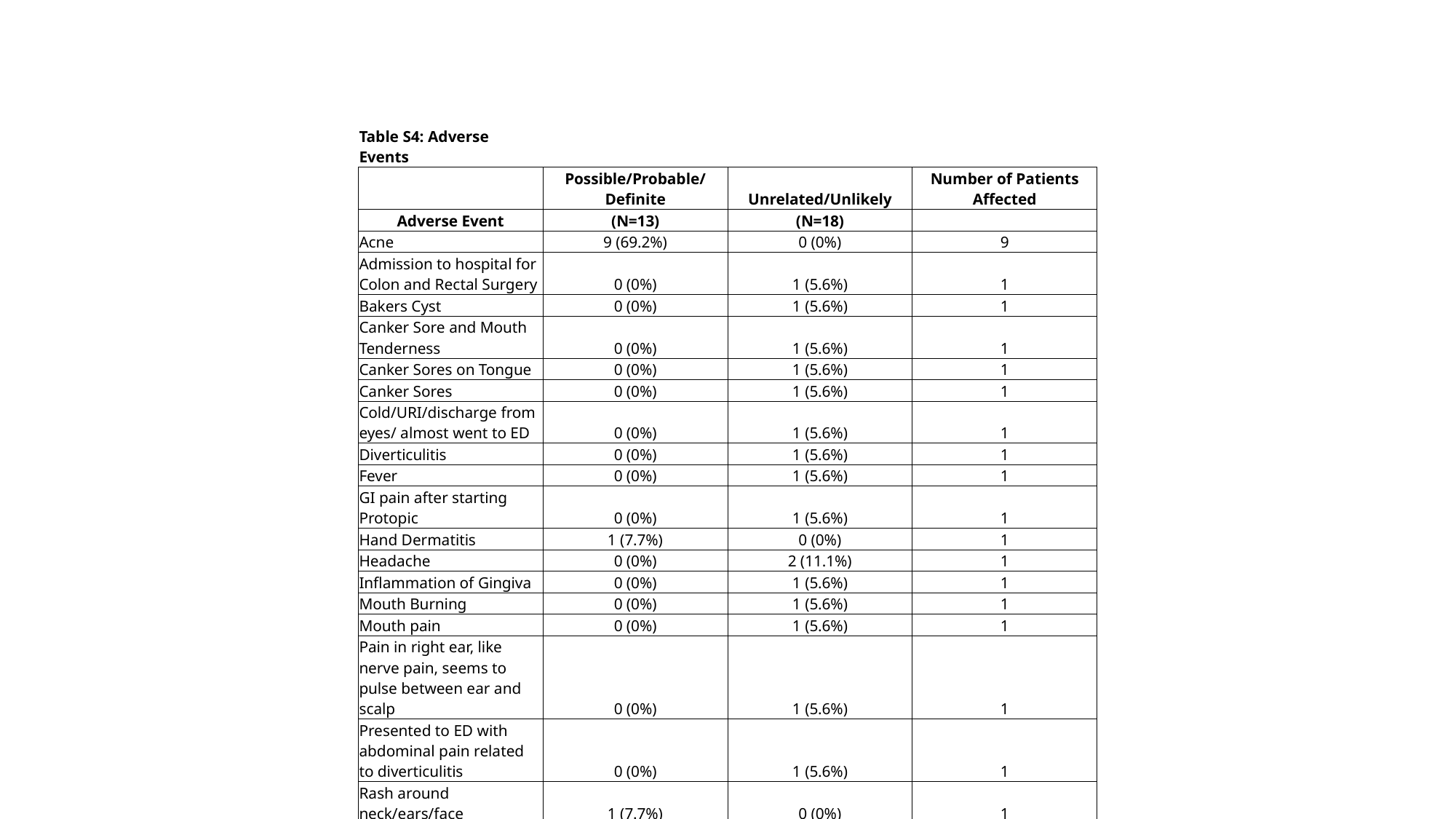

| Table S4: Adverse Events | | | |
| --- | --- | --- | --- |
| | Possible/Probable/Definite | Unrelated/Unlikely | Number of Patients Affected |
| Adverse Event | (N=13) | (N=18) | |
| Acne | 9 (69.2%) | 0 (0%) | 9 |
| Admission to hospital for Colon and Rectal Surgery | 0 (0%) | 1 (5.6%) | 1 |
| Bakers Cyst | 0 (0%) | 1 (5.6%) | 1 |
| Canker Sore and Mouth Tenderness | 0 (0%) | 1 (5.6%) | 1 |
| Canker Sores on Tongue | 0 (0%) | 1 (5.6%) | 1 |
| Canker Sores | 0 (0%) | 1 (5.6%) | 1 |
| Cold/URI/discharge from eyes/ almost went to ED | 0 (0%) | 1 (5.6%) | 1 |
| Diverticulitis | 0 (0%) | 1 (5.6%) | 1 |
| Fever | 0 (0%) | 1 (5.6%) | 1 |
| GI pain after starting Protopic | 0 (0%) | 1 (5.6%) | 1 |
| Hand Dermatitis | 1 (7.7%) | 0 (0%) | 1 |
| Headache | 0 (0%) | 2 (11.1%) | 1 |
| Inflammation of Gingiva | 0 (0%) | 1 (5.6%) | 1 |
| Mouth Burning | 0 (0%) | 1 (5.6%) | 1 |
| Mouth pain | 0 (0%) | 1 (5.6%) | 1 |
| Pain in right ear, like nerve pain, seems to pulse between ear and scalp | 0 (0%) | 1 (5.6%) | 1 |
| Presented to ED with abdominal pain related to diverticulitis | 0 (0%) | 1 (5.6%) | 1 |
| Rash around neck/ears/face | 1 (7.7%) | 0 (0%) | 1 |
| Rash on neck | 0 (0%) | 1 (5.6%) | 1 |
| Rash/acne | 1 (7.7%) | 0 (0%) | 1 |
| Sore throat/lymph node area/sickness | 0 (0%) | 1 (5.6%) | 1 |
| Virus (cold/flu) | 1 (7.7%) | 0 (0%) | 1 |

### Slide 9
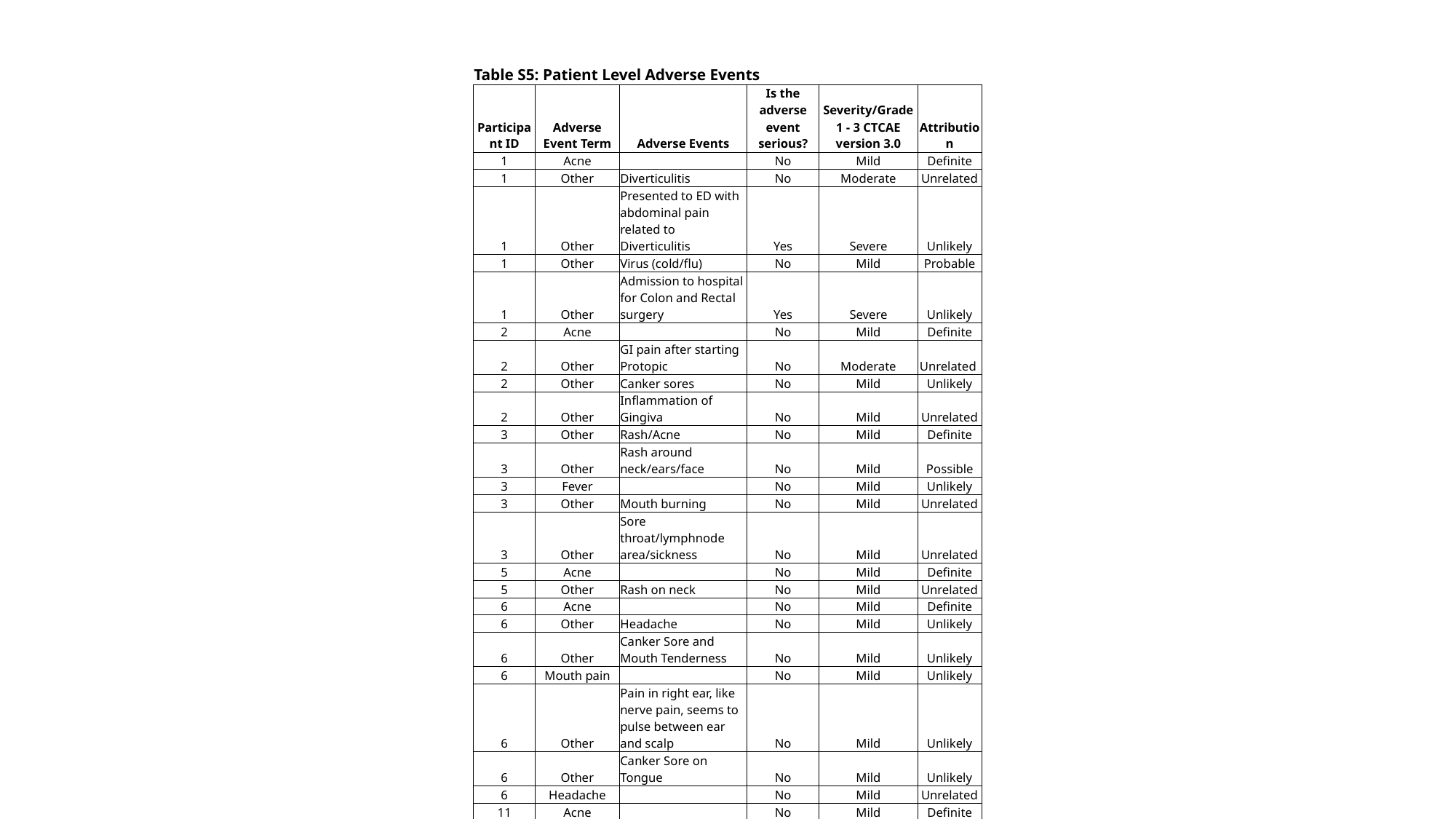

| Table S5: Patient Level Adverse Events | | | | | |
| --- | --- | --- | --- | --- | --- |
| Participant ID | Adverse Event Term | Adverse Events | Is the adverse event serious? | Severity/Grade 1 - 3 CTCAE version 3.0 | Attribution |
| 1 | Acne | | No | Mild | Definite |
| 1 | Other | Diverticulitis | No | Moderate | Unrelated |
| 1 | Other | Presented to ED with abdominal pain related to Diverticulitis | Yes | Severe | Unlikely |
| 1 | Other | Virus (cold/flu) | No | Mild | Probable |
| 1 | Other | Admission to hospital for Colon and Rectal surgery | Yes | Severe | Unlikely |
| 2 | Acne | | No | Mild | Definite |
| 2 | Other | GI pain after starting Protopic | No | Moderate | Unrelated |
| 2 | Other | Canker sores | No | Mild | Unlikely |
| 2 | Other | Inflammation of Gingiva | No | Mild | Unrelated |
| 3 | Other | Rash/Acne | No | Mild | Definite |
| 3 | Other | Rash around neck/ears/face | No | Mild | Possible |
| 3 | Fever | | No | Mild | Unlikely |
| 3 | Other | Mouth burning | No | Mild | Unrelated |
| 3 | Other | Sore throat/lymphnode area/sickness | No | Mild | Unrelated |
| 5 | Acne | | No | Mild | Definite |
| 5 | Other | Rash on neck | No | Mild | Unrelated |
| 6 | Acne | | No | Mild | Definite |
| 6 | Other | Headache | No | Mild | Unlikely |
| 6 | Other | Canker Sore and Mouth Tenderness | No | Mild | Unlikely |
| 6 | Mouth pain | | No | Mild | Unlikely |
| 6 | Other | Pain in right ear, like nerve pain, seems to pulse between ear and scalp | No | Mild | Unlikely |
| 6 | Other | Canker Sore on Tongue | No | Mild | Unlikely |
| 6 | Headache | | No | Mild | Unrelated |
| 11 | Acne | | No | Mild | Definite |
| 12 | Other | Cold/URI/discharge from eyes, almost went to ER | No | Moderate | Unlikely |
| 12 | Acne | | No | Mild | Probable |
| 13 | Acne | | No | Mild | Definite |
| 14 | Acne | | No | Mild | Definite |
| 15 | Acne | | No | Mild | Definite |
| 15 | Other | Bakers Cyst | No | Moderate | Unrelated |
| 15 | Other | Hand Dermatitis | No | Mild | Possible |
